# Evaluation of a disease-specific mHealth-based exercise self-tracking measure

**DOI:** 10.1101/2022.05.16.22275170

**Authors:** Ipek Ensari, Emma Horan, Noémie Elhadad, Suzanne R. Bakken

## Abstract

**Objectives:** This study investigates the concurrent and construct validity of a brief, customizable exercise self-tracking item from a research mHealth App (“Phendo”) for use as a measure of day-level and habitual exercise behavior in endometriosis.

**Study Sample:** Study 1 included 52 participants who were recruited online and provided data for up to 14 days. Study 2 included 359 Phendo users who had retrospectively-collected data on the study measures.

**Methods:** In Study 1, we evaluated the responses on the self-tracking exercise item as estimates of day-level moderate-to-vigorous intensity exercise (MVE) and total step counts. Comparison measures included recall-based MVE minutes and accelerometry-based step counts, which were self-reported through daily surveys. In Study 2, we derived a measure of habitual exercise using each individual’s longitudinal self-tracked responses. We assessed its concurrent validity using the Nurses’ Health Study II Physical Activity Scale (NHS-II) as the comparison measure. We assessed its discriminant validity through known-group differences analysis where the sample was dichotomized based on Health Survey Short Form-36 (SF-36) and body mass index (BMI).

**Data Analysis:** We assessed bivariate associations between the scores on the self-tracking and comparison measures using Kendall’s rank correlations. We estimated daily MVE and step counts (Study 1), and weekly exercise (Study 2) from the self-tracking item scores through adjusted linear and polynomial regression models. We used t-tests and linear regression to conduct known-group differences analyses.

**Results:** In Study 1, self-tracked exercise responses were moderately correlated with survey based MVE and step counts. Regression analyses indicated that overall exercise responses were associated with ∼17 minutes of MVE for the average participant (B=16.09, t=3.11, p=0.045). Self-tracked aerobic-type exercise was a stronger predictor of MVE minutes and step counts (B=27.561, t=5.561, p<0.0001). In Study 2, each self-tracked exercise instance corresponded to ∼19 minutes of exercise per week on the NHS-II Scale (B=19.80, t=2.1, p=0.028). Finally, there were statistically significant differences between the groups dichotomized based on SF-36 subscale scores and BMI.

**Conclusion:** This study presents preliminary evidence on the concurrent and discriminant validity of a brief mHealth App measure for exercise self-tracking among individuals with endometriosis. These findings have implications in the context of large-scale studies that involve monitoring a diverse group of participants over long durations of time, as well as engaging and retaining research participants.

## Introduction

The emergence of mobile and wearable health (mHealth) technologies is rapidly expanding their use in research and clinical settings,[1-3] and engaging patients in self-management and monitoring of their conditions.[4, 5] mHealth-based digital measures that allow daily self-tracking and event-based reporting constitute a promising method for administering frequent, low-burden assessments in natural settings to gain important information about patients’ health status and make clinical decisions. Moreover, such measures are often shorter (e.g., single-item) compared to traditional self-report measures and user-customizable, i.e., the same construct could be measured through a different set of items by the participants. Thus, mHealth-based daily self-report data on health outcomes circumvent several limitations of data from traditional self-report measures, including limited observation period, recall bias,[6] and lack of granularity of the patient experience and health status.[7, 8]

Yet, there is a scarcity of studies evaluating the validity and reliability of mHealth-based self-tracking measures.[1, 2, 9-11] This has been identified as a high priority research area for mHealth evidence generation,[12] and is critical to advance mHealth methods and have meaningful impact toward improving public health.[1, 2, 9-11] Moreover, investigating the validity of customizable, disease-specific self-tracking measures is necessary to improve their application and understand their relevance to the specific patient population. Accordingly, there is a need to assess the validity of mHealth-based measures designed for daily self-monitoring of health behaviors by individuals with chronic conditions toward symptom self-management.

Herein, we investigate the validity of a mHealth-based exercise self-tracking measure designed for individuals with endometriosis.[13, 14] Endometriosis is a systemic, estrogen-dependent inflammatory condition characterized primarily by chronic pelvic and abdominal pain.[15, 16] It has high societal burden due to loss of work productivity and impact on quality of life (QoL). [17-20] There are substantial between-patient heterogeneity and day-to-day fluctuations in its symptomology,[5, 21] and existing medical therapies have limited efficacy, often confounded by side effects.[22] Moreover, evidence suggests that individuals with endometriosis are interested in daily self-monitoring of symptoms and various health behaviors daily for better disease self-management. These factors collectively make mHealth methods particularly valuable in the context of endometriosis for capturing patient-reported outcomes and health behaviors over time, and provide further motivation for undertaking this work.

We focus on exercise behavior (i.e., leisure-time physical activity (PA) that is done repeatedly with the end goal to improve fitness) as it constitutes an important component of health and disease management.[23, 24] Both acute (i.e., single bout/session) and chronic (i.e., repeated bouts/sessions over time) exercise have been demonstrated to improve numerous disease outcomes and related symptoms.[23, 25-29] Previous studies relying on recall-based survey data suggested that individuals with endometriosis are less likely to engage in adequate amounts of regular exercise, which in return is a risk factor for exacerbating disease progression. On the other hand, there is some evidence from mHealth-based daily self-tracked data that individuals with endometriosis engage in a variety of exercise modalities. It is possible that their needs with regards to a self-tracking measure of exercise may differ from those of other populations. Assessing the validity of a mHealth-based daily self-tracking exercise measure designed for individuals with endometriosis is a starting point for delineating these gaps in the literature.

In this work, we investigate the concurrent[30] and discriminant[31] validity of a mHealth-based exercise self-tracking item from an observational research mHealth App (“Phendo”) for endometriosis.[13, 14] Phendo was previously developed using patient-centered participatory design, through qualitative (focus groups, interviews) and quantitative research (surveys, coded content analysis) with participants with endometriosis, described in detail elsewhere.[13, 14] This technique for developing patient-reported outcome measures has been suggested to enhance content validity and relevance of the measure to the target demographic, thus providing a more comprehensive and accurate representation of the disease experience and impact.[32-35]

Our preliminary analyses toward validation of Phendo’s exercise self-tracking item considers different time frames. First, we evaluate it as a day-level measure and assess the concurrent validity of the responses as estimates of overall and different modalities of exercise (Study 1). We then use each individual’s longitudinal responses on the self-tracking item to derive a measure of habitual exercise (i.e., average patterns over the long term) and evaluate its concurrent and discriminant validity (Study 2).

## Study 1 – Day level measure evaluation

### Methods

#### Sample and Recruitment

All procedures were approved by Columbia University Irving Medical Center Institutional Review Board and all participants provided informed consent (#AAAQ9812). We recruited participants through advertisements on Phendo’s social media accounts (citizenendo.org, Twitter, Instagram, medium blog) in November 2018, where the participation opportunity was advertised as a voluntary (i.e., unpaid), 2-week study aimed to better understand PA and exercise habits among individuals with endometriosis. Participants were instructed to maintain their usual levels of PA and exercise during this 2-week period. Eligibility criteria included individuals with an endometriosis diagnosis, interest in tracking/monitoring PA and exercise behavior, willingness to respond to daily mobile surveys on PA and exercise behavior, and self-track disease symptoms and exercise behavior in Phendo every day for 14 days. Though the study was advertised on the App’s social media pages, participation was not restricted to current Phendo users.

#### Study Measures and Variables

##### Self-tracking exercise measures

We evaluated 3 day-level outcome measures derived from the self-tracking exercise item: Overall (any modality/intensity), aerobic-type, and multimodal/anaerobic-type exercise. Exercise is tracked at the day level within the Phendo App through a root question “Did you exercise today? (Yes/No)”. Upon selecting a “Yes”, users can further create customized exercise tracking items by adding details (e.g., modality, intensity, duration) within this question. These customized tracking items can be saved within the App for later use to eliminate re-entry every time the user wants to track any activity they regularly do. Responses to the root question were used as a binary measure of overall day-level exercise for comparison to other measures from the daily surveys for assessment of concurrency. We extracted the free-text data from the customized item responses on modality to derive the 2 other binary variables including: 1) Aerobic type (e.g., walking, running, jogging, etc.), and 2) Anaerobic/Multimodal type (e.g., yoga, Pilates, calisthenics, strength, etc.). Walking- and most aerobic-type exercises are considered to be of moderate or vigorous intensity according to the Compendium of PA [36], which is a standardized way of categorizing different PA types for measurement of PA behavior in research. Moreover, most aerobic type exercises reported by the participants were step-based (e.g., jogging, running, elliptical and stair machines). We expected the aerobic exercise responses to correlate most strongly with those on the comparison MVE and step count measure. Accordingly, we hypothesized that the anaerobic/multimodal exercise variable would be relatively weakly correlated with the comparison measure responses.

##### Comparison measures

As comparison measures, we used self-reported daily step counts and MVE minutes from body-worn accelerometers and the 1-item Exercise Vital Sign (EVS).[37] EVS provides an estimate of leisure-time PA (i.e., exercise) behavior and has been demonstrated to discriminate patients with differing activity levels based on demographics and health status. We adapted this item for day-level administration, i.e., “How many minutes of physical activity *that is enough to raise your breathing rate* did you do today? This may include sport, exercise, brisk walking or cycling for recreation or to get to and from places but should not include housework or physical activity that may be part of your job’. Comparison measures were administered every day through daily surveys sent through Qualtrics to participants’ email addresses.

We used the MVE responses to create 3 comparison measures: 1) As total daily minutes, 2) Categorized into 2 levels: less than or at least 30 minutes (MVE 2-level category), and 3) Categorized into 3 levels: less than 30 minutes, 30-149 minutes, at least 150 minutes (MVE 3-level category). Given the step counts obtained through body-worn trackers are not limited to just periods of leisure-time MVE, they capture all intensities and types of PA throughout the day. Accordingly, we used the step counts outcome as a comparison measure of overall day-level PA, and MVE responses as a comparison measure of overall day-level MVE (i.e., leisure-time PA of at least moderate intensity).

#### Data Analysis

We describe the demographic characteristics of the study sample using means and standard deviations (SDs) and ranges for all variables. For a comprehensive assessment of concurrent validity, we assessed several metrics of association between the Phendo items and the daily survey items on MVE and Steps. First, we computed Kendall’s rank correlation tau coefficients (*τ*) to quantify the magnitude of bivariate associations between the responses from the Phendo exercise items and those from the daily surveys. Next, we assessed the magnitude of associations while adjusting for number of tracked days to partial out the potential variance brought in by an individual’s tracking habits and/or number of days of data. This was done by conducting separate linear regression models with daily step counts and MVE minutes as the outcomes. In all of the models, the outcome was regressed on one of the 3 types of self-tracked exercise responses (i.e., any-, aerobic-, multimodal type exercise) and adjusted for the number of tracked days.

## Study 1 Results

### Sample Descriptive Statistics

Fifty-two participants initially expressed interest in participating, and completed informed consent and the first day of Qualtrics survey questions. Of those, 39 participants provided at least 2 days of survey data, and 31 provided at least 3 days of data (Mean (SD)=6.44 (5.04) days, Range=1-14). In total, 335 person-level days of data from 52 participants were available for analysis. Twenty-one participants reported wearing an activity tracker on 1 or more days during the 14-day period (Mean (SD) = 5.49 (4.65) days), providing 146 person-level days of tracker-based data. Descriptive summary statistics for the Study 1 sample on demographics, daily survey responses, and the self-tracked exercise reports are provided in Table 1.

**Table 1.** Sample summary statistics on age, body mass index, daily survey and self-tracking measures for the Study 1 sample (N=52).

| Sample Demographics | Mean (SD)/ Frequency(%) | Range |
| --- | --- | --- |
| Age | 30.90 (6.91) | 18-53 (Median=31.50) |
| Body Mass Index | 25.02 (6.63) | 17.08-44.2<br>(Median=22.74) |
| <u>Race/Ethnicity</u> |  |  |
| Caucasian White | 44 (84.6%) |  |
| Asian | 1 (1.9%) |  |
| Non-Hispanic Black | 1 (1.9%) |  |
| Hispanic | 3 (5.7%) |  |
| Other | 3 (5.7%) |  |
| <u>Education</u> |  |  |
| College or higher | 34 (65.3%) |  |
| Some College | 15 (28.8%) |  |
| High School or less | 3 (5.7%) |  |
| <u>Employment</u> |  |  |
| Employed | 35 (67.3%) |  |
| Not Employed | 9 (17.3%) |  |
| Student | 8 (15.3%) |  |
| **Daily survey measure (N)** | **Mean (SD)** | **Range** |
| Steps (21)<br>(146 person-level days) | 6779.75 (4444.99) | 12-18,858 |
| MVE minutes (31)<br>(333 person-level days) | 31.96 (35.11) | 0-240 |
| <u>Time of MVE</u> |  |  |
| Between wake time to Noon (31) | 137 (40.8%) |  |
| Noon to 6pm (36) | 188 (56.1%) | N/A |
| 6pm to bedtime (24) | 67 (20.0%) |  |
| Tracker wear time (21) | 16.64 (6.4) | 3-24 (146) |
| **Phendo exercise measure** | **Frequency (%)** |  |
| <u>"Did you do any exercise today?"</u> |  |  |
| Yes (25) | 84 (25.0%) |  |
| No (22) | 109 (32.5%) |  |
| NA (42) | 142 (42.3%) |  |
| <u>Modality</u> |  |  |
| Walking (14) | 59 (17.6%) |  |
| Aerobic (22) | 78 (23.2%) |  |
| Strength (1) | 3 (0.8%) |  |
| Multimodal (12) | 25 (7.4%) |  |

### Concurrent validity

Results from the bivariate rank correlations are provided in Table 2. As an overall daily exercise measure, responses on the self-tracking item were moderately correlated with daily survey-based MVE outcomes and step counts (*τ* = 0.256 and *τ* = 0.294, respectively; p<0.0001, corresponding to Fisher’s z scores of 0.41 and 0.48). As expected, aerobic-type exercise responses were more strongly correlated than anaerobic/multimodal exercise responses with the MVE and step count outcomes.

**Table 2.** Results of the Kendall’s rank correlations between responses from the self-tracked exercise items and daily survey MVE and step counts.

|  | **MVE Mins** | **MVE 2-level category** | **MVE 3-level category** | **Total Steps** |
| --- | --- | --- | --- | --- |
| **Self-tracked exercise (any)** | $\tau = 0.256$,<br>$z = 4.171$,<br>$p < 0.0001$ | $\tau = 0.252$,<br>$z = 3.502$,<br>$p = 0.0004$ | $\tau = 0.252$,<br>$z = 3.520$,<br>$p = 0.0004$ | $\tau = 0.294$,<br>$z = 3.232$,<br>$p = 0.001229$ |
| **Self-tracked aerobic exercise** | $\tau = 0.397$,<br>$z = 6.461$,<br>$p<0.00001$ | $\tau = 0.3382$,<br>$z = 4.6863$,<br>$p<0.00001$ | $\tau = 0.336$,<br>$z = 4.692$,<br>$p<0.00001$ | $\tau = 0.361$,<br>$z = 4.033$,<br>$p < 0.0001$ |
| **Self-tracked multimodal exercise** | $\tau = 0.057,$<br>$z = 0.9406,$<br>$p = 0.346$ | $\tau = 0.118,$<br>$z = 1.63,$<br>$p = 0.101$ | $\tau = 0.123,$<br>$z = 1.725,$<br>$p = 0.08446$ | $\tau = 0.159,$<br>$z = 1.790,$<br>$p = 0.0733$ |

Results of the adjusted and unadjusted regression models estimating MVE minutes are provided in Table 3. When the overall daily exercise measure was used as the predictor, point estimates indicated that each self-tracked exercise instance was associated with an additional ∼16 minutes of MVE for the average participant (B=16.09, t=3.11, p=0.045). However, aerobic-type exercise was a stronger predictor where each self-tracked aerobic exercise instance was associated with an additional ∼14 minutes of MVE for a total of ∼28 minutes (B=27.561, t=5.561, p<0.0001). In contrast, multimodal exercise was not a significant predictor of MVE mins (See Table 3). The results from the models estimating step counts are provided in Table 4, which indicated that each self-tracked exercise instance was associated with an additional 3,226 steps on a given day. For self-tracked aerobic exercise responses, the point estimates were larger by ∼500 steps (B=3766.5, t=5.12, p<0.0001), whereas multimodal exercise responses were similar to the overall exercise responses (B=3240.2, t=2.231, p=0.028).

**Table 3.** Results for the 3 separate linear regression models estimating daily survey responses on MVE minutes from self-tracked exercise responses (i.e., any-, aerobic-, multimodal-exercise).

| Model Term | B coefficient (SE) | t-value | P value |  |
| --- | --- | --- | --- | --- |
| **Outcome= MVE Mins** |  |  |  |  |
| Intercept | 20.712 (10.273) | 2.016 | 0.045 |  |
| Self-tracked exercise (any) | 16.091 (5.166) | 3.115 | 0.045 |  |
| Tracked days | 0.513 (0.799) | 0.643 | 0.521 | F(190)=4.917, p=0.0082 |
| Intercept | 20.010 (9.49) | 2.108 | 0.036 |  |
| Self-tracked aerobic exercise | 27.561 (4.940) | 5.579 | <0.0001 |  |
| Tracked days | 0.224 (0.756) | 0.297 | 0.766 | F(190)=15.64, p <0.00001 |
| Intercept | 26.537 (10.220) | 2.596 | 0.010 |  |
| Self-tracked multimodal exercise | 13.195 (7.762) | 1.693 | 0.092 |  |
| Tracked days | 0.469 (0.817) | 0.574 | 0.566 | F(190)=1.496, p=0.226 |

**Table 4.**
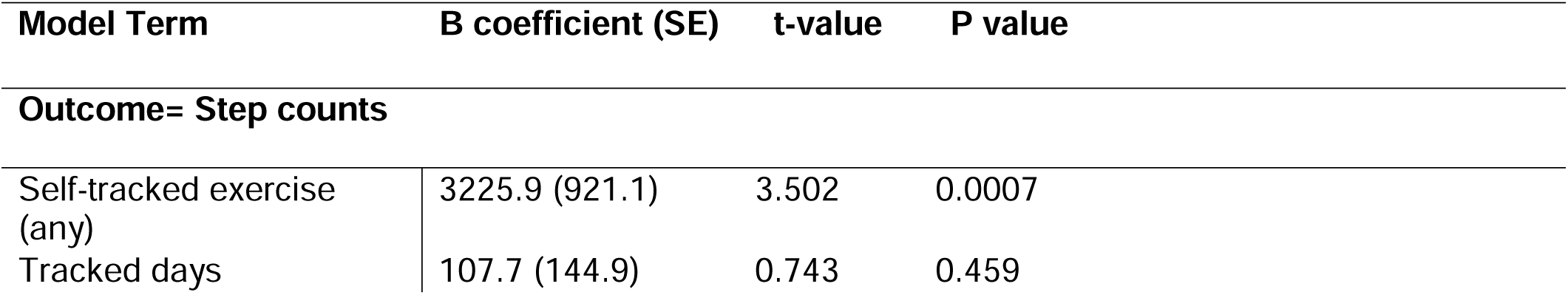
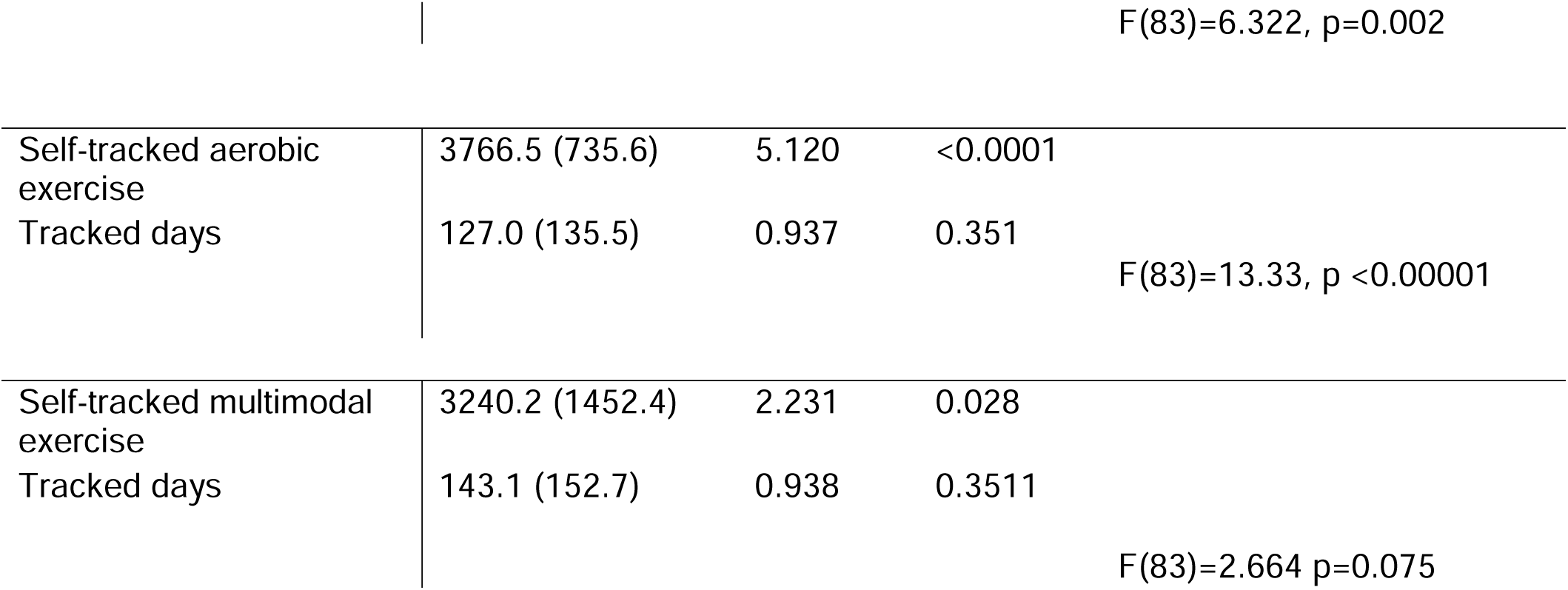
Results for the 3 separate linear regression models estimating daily survey based step counts from self-tracked exercise outcomes (i.e. any-, aerobic-, multimodal exercise).

| Model Term | B coefficient (SE) | t-value | P value |
| --- | --- | --- | --- |
| **Outcome= Step counts** |  |  |  |
| Self-tracked exercise (any) | 3225.9 (921.1) | 3.502 | 0.0007 |
| Tracked days | 107.7 (144.9) | 0.743 | 0.459 |

|  |  |  |  |  |
| --- | --- | --- | --- | --- |
|  |  |  |  | F(83)=6.322, p=0.002 |
| Self-tracked aerobic exercise | 3766.5 (735.6) | 5.120 | <0.0001 |  |
| Tracked days | 127.0 (135.5) | 0.937 | 0.351 | F(83)=13.33, p <0.00001 |
| Self-tracked multimodal exercise | 3240.2 (1452.4) | 2.231 | 0.028 |  |
| Tracked days | 143.1 (152.7) | 0.938 | 0.3511 | F(83)=2.664 p=0.075 |

## Study 2 – Evaluation of the habitual exercise measure

### Methods

#### Study Sample

All procedures were approved by Columbia University Irving Medical Center Institutional Review Board and all participants provided informed consent (#AAAQ9812). Analyses for Study 2 were conducted with retrospective data collected through Phendo between November 2016 and April 2020. Details of recruitment, enrollment, informed consent are described elsewhere.[5, 38] Briefly, participants consisted of a subset of Phendo App users who had longitudinal self-tracked exercise data for derivation of a habitual exercise measure and self-reported a surgery-, clinician-, or suspected (i.e., self-) diagnosis of endometriosis within their App profiles. This was an a priori decision based on the focus of the study (i.e., exercise behavior) and our previous findings indicating no substantial differences in exercise patterns between those with self-diagnosed vs formally-diagnosed endometriosis. All participants provided informed consent prior to data collection and all Phendo users contribute data with the acknowledgement that their de-identified data can be used for research purposes.

#### Study Measures and Variables

##### Phendo habitual exercise measure

For each participant, we computed their mean weekly exercise frequency (i.e., habitual exercise) by summing their self-tracked exercise reports per week across their entire range of days of data and then dividing this number by the total number of weeks of data they had. We used the responses to the same root exercise question in Phendo described in Study 1 Methods to compute this habitual exercise proxy measure.

##### Comparison measures

We used the 8-item Nurses’ Health Study II PA (NHS-II) Scale [39] included within the World Endometriosis Research Foundation (WERF) Endometriosis Patient Questionnaire (EPQ-S)[40, 41] as the comparison instrument.MThe NHS-II Scale asks the respondent to report the typical weekly durations spent in major recreational PA categories (i.e., walking, running, lap swimming, jogging, bicycling, tennis, calisthenics, other aerobic recreation) in the past 12 months. We added these durations to obtain total weekly raw minutes and metabolic equivalent minutes (MET-mins) of recreational PA (i.e., exercise). MET-mins of exercise is computed by multiplying the duration of each activity by its MET intensity level based on the Compendium of PA.[36] The MET intensity reflects the associated metabolic rate for a specific activity divided by a standard resting metabolic rate. One MET-min is roughly equivalent to 1 kcal/min for a 60-kg person. METs are most commonly used to categorize PAs based on intensity where, moderate intensity is defined as 3–6 METs, moderate-to-vigorous intensity as >3 METs, and vigorous intensity as >6 METs.[42, 43]

Finally, we used the Physical Function and Energy subscales of the 36-Item Short Form Health Survey (SF-36)[44] and body mass index (BMI) as the dichotomization variables to assess discriminant validity via known-group differences analyses. The SF-36 is a set of self-reported functioning and well-being measures developed for the Medical Outcomes Study, a large-scale study of how patients fare with health care in the United States.[45] The subscale scores are computed by summing the weighted scores from each item and converted to standardized T-scores. The T-scores range from 0 to 100 where higher scores indicate less disability and 50 represents the population normative mean.

#### Data Analysis

##### Concurrent validity

The EPQ-S is included within the Phendo App for participants to complete if and when they would like. Thus, complete data were available for a subset of the Phendo users and there was variability across participants in terms of the when they completed it in relation to the time of their self-tracking. We included all available data for analysis and adjusted the models for time duration between self-tracking in Phendo and NHS-II Scale to account for any potential variance brought in by this time interval. We first conducted Kendall’s rank correlations between the person-level median habitual exercise scores and the NHS-II weekly exercise outcomes (weekly raw minutes and MET-mins). Next, we conducted separate linear regression models where the NHS-II outcomes were each regressed on the habitual exercise scores, and further adjusted for duration between self-tracking and survey completion. We used the median values for the habitual exercise outcome in the regression models for interpretability. Given that the habitual exercise outcome is based on weekly frequency and does not take into account intensity of the exercise, we expected to be able to capture its relationship with the raw minutes of exercise from NHS-II through a linear model. On the other hand, we hypothesized a possible non-linear relationship with the MET-mins of exercise (which takes into account intensity), and tested for this through higher-order polynomial regressions.

##### Discriminant Validity

We evaluated the construct validity of the habitual exercise measure by assessing its discriminant validity via known-group differences analysis. In this type of analysis, the sample is categorized into groups based on selected person-level variables that are hypothesized to be associated with different scores on the measure of interest (i.e. habitual exercise).[46] Accordingly, we used the SF-36 subscales of Physical Function and Energy, and BMI category (i.e., <18.5, 18.5-24.99, 25-29.99, >=30) as the grouping variables. We hypothesized that those with lower scores on the SF-36 subscales and BMIs of 30 or greater (based on previous research on BMI and PA [47]), would be associated with significantly lower habitual exercise levels. We used the population normative means on the SF-36 as the cut-off score (i.e., >50 vs _≤_50) to create the groups, and conducted independent samples t-tests or linear regression where appropriate to compare habitual exercise levels across the groups.

## Study 2 Results

### Sample characteristics

Participants (N=359) had on average 54.3 days of data available for analysis (SD=64.3, Range=11-395). Demographics for the participants are provided in Table 5. Participants collectively represented 27 countries (N=156 residing in the United States), and ages across the adult reproductive span (14.3-49.2 years). Overall sample mean for habitual exercise derived from Phendo’s self-tracking item was 1.45 times per week (SD=1.48, Range=1-7, Median=1). Of note, 176 (49.0%) participants had a mean weekly exercise frequency of fewer than once per week. Average weekly minutes of exercise based on the NHS-II was 174.95 (SD=280.12, Range=0-2790, Median=72). Average weekly MET-mins was 967.9 (SD=1822.25, Range=0-20265, Median=316.86).

**Table 5.**
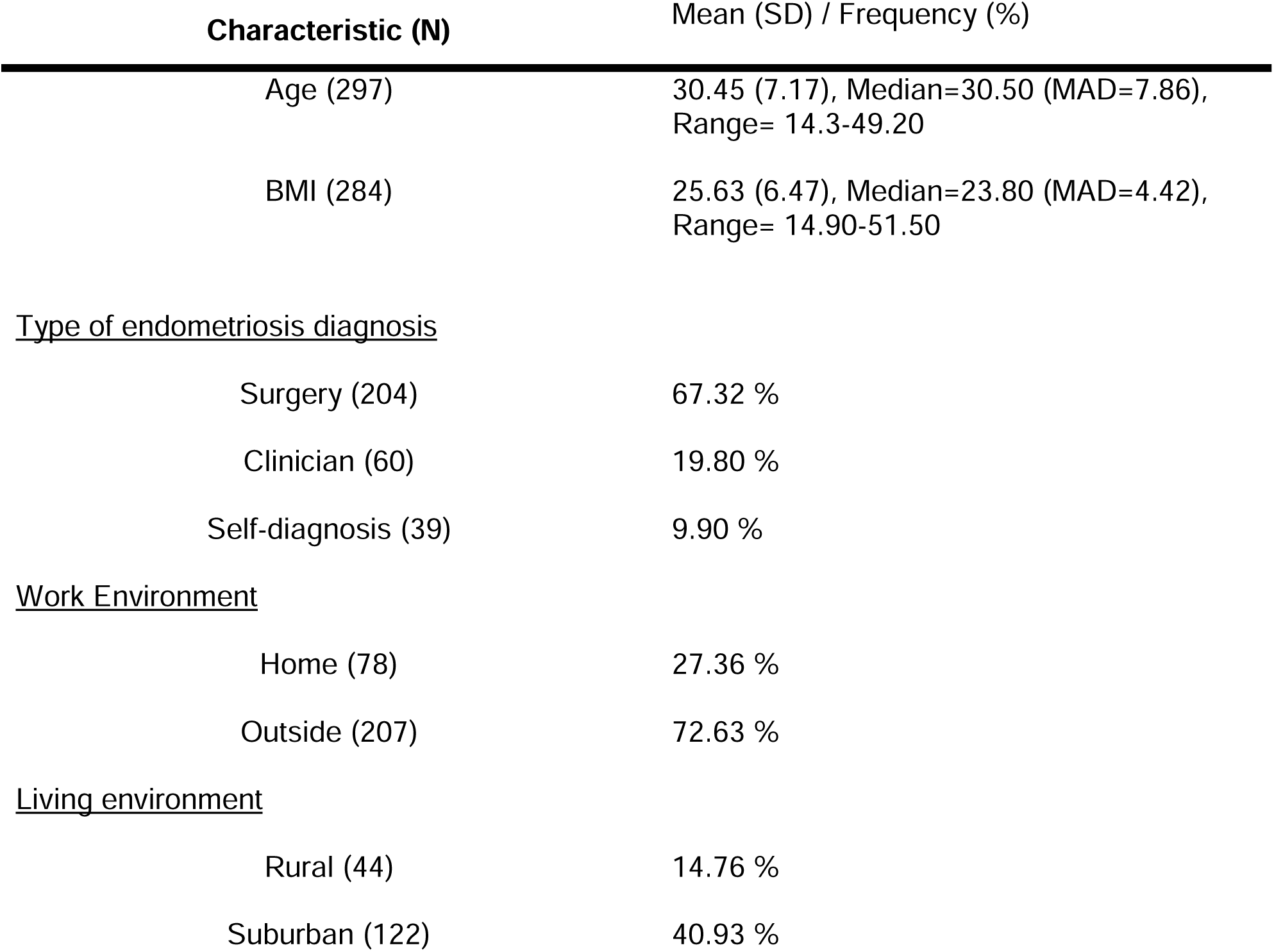
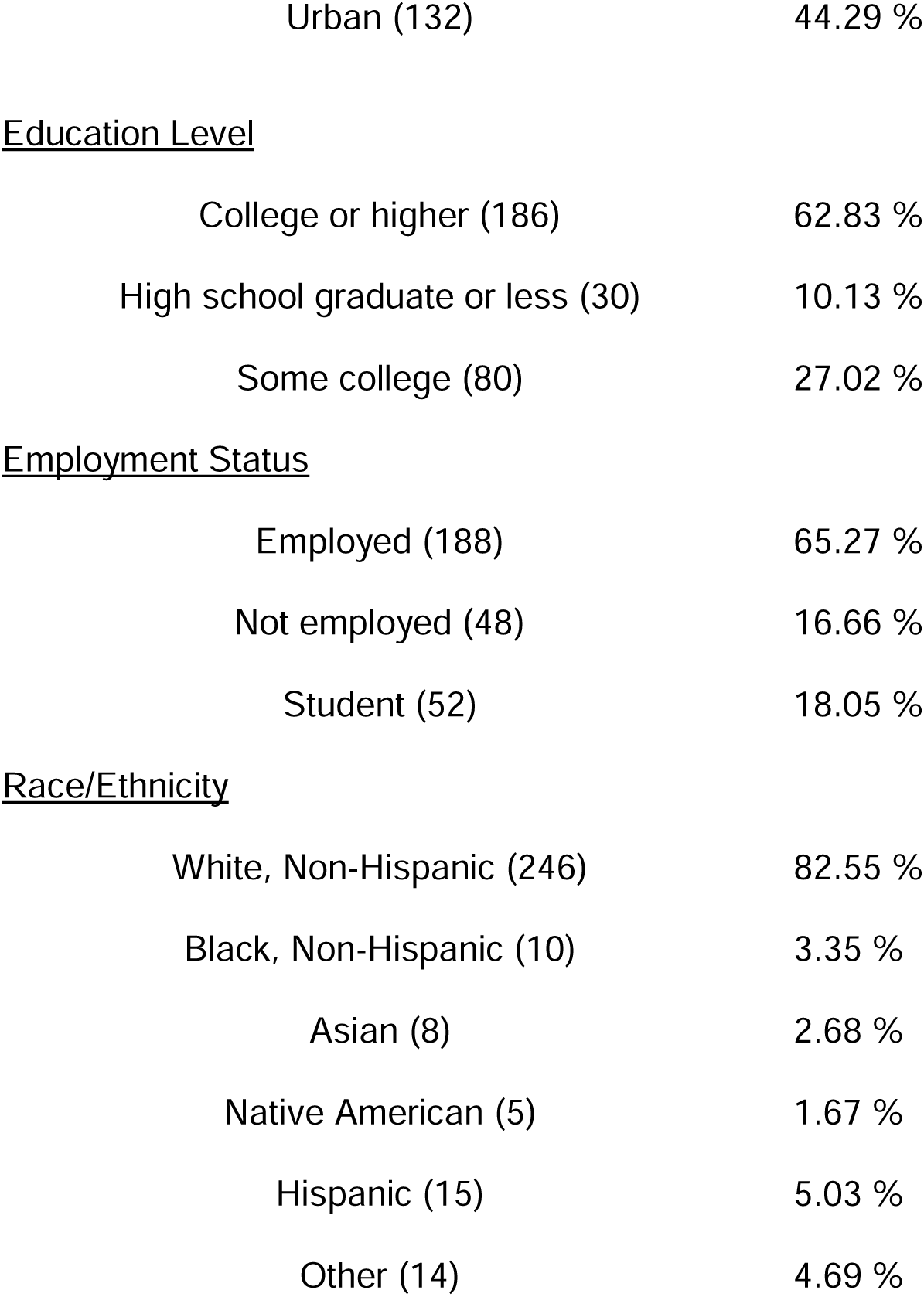
Study 2 Sample characteristics (N=359).

| Characteristic (N) | Mean (SD) / Frequency (%) |
| --- | --- |
| Age (297) | 30.45 (7.17), Median=30.50 (MAD=7.86), Range= 14.3-49.20 |
| BMI (284) | 25.63 (6.47), Median=23.80 (MAD=4.42), Range= 14.90-51.50 |
| <u>Type of endometriosis diagnosis</u> |  |
| Surgery (204) | 67.32 % |
| Clinician (60) | 19.80 % |
| Self-diagnosis (39) | 9.90 % |
| <u>Work Environment</u> |  |
| Home (78) | 27.36 % |
| Outside (207) | 72.63 % |
| <u>Living environment</u> |  |
| Rural (44) | 14.76 % |
| Suburban (122) | 40.93 % |
| Urban (132) | 44.29 % |
| Education Level |  |
| College or higher (186) | 62.83 % |
| High school graduate or less (30) | 10.13 % |
| Some college (80) | 27.02 % |
| Employment Status |  |
| Employed (188) | 65.27 % |
| Not employed (48) | 16.66 % |
| Student (52) | 18.05 % |
| Race/Ethnicity |  |
| White, Non-Hispanic (246) | 82.55 % |
| Black, Non-Hispanic (10) | 3.35 % |
| Asian (8) | 2.68 % |
| Native American (5) | 1.67 % |
| Hispanic (15) | 5.03 % |
| Other (14) | 4.69 % |

### Concurrent Validity

Habitual exercise outcome was moderately correlated with NHS-II scores for both raw minutes (i.e., *τ* = 0.18, z = 4.52, p<0.0001) and MET-mins of exercise (*τ* = 0.17, z = 4.39, p<0.0001). All results from the regression models are provided in Table 6. In both the adjusted and unadjusted models estimating NHS-II weekly raw exercise minutes, the B coefficients for self-tracking based habitual exercise variable were statistically significant (p<0.05 for both point estimates and the overall model fit statistics). The results indicated that, each self-tracked exercise instance in a typical week was associated with an additional ∼19 minutes of exercise on the NHS-II (B=19.80 and B=18.73 in the unadjusted and adjusted models, respectively).

**Table 6.**
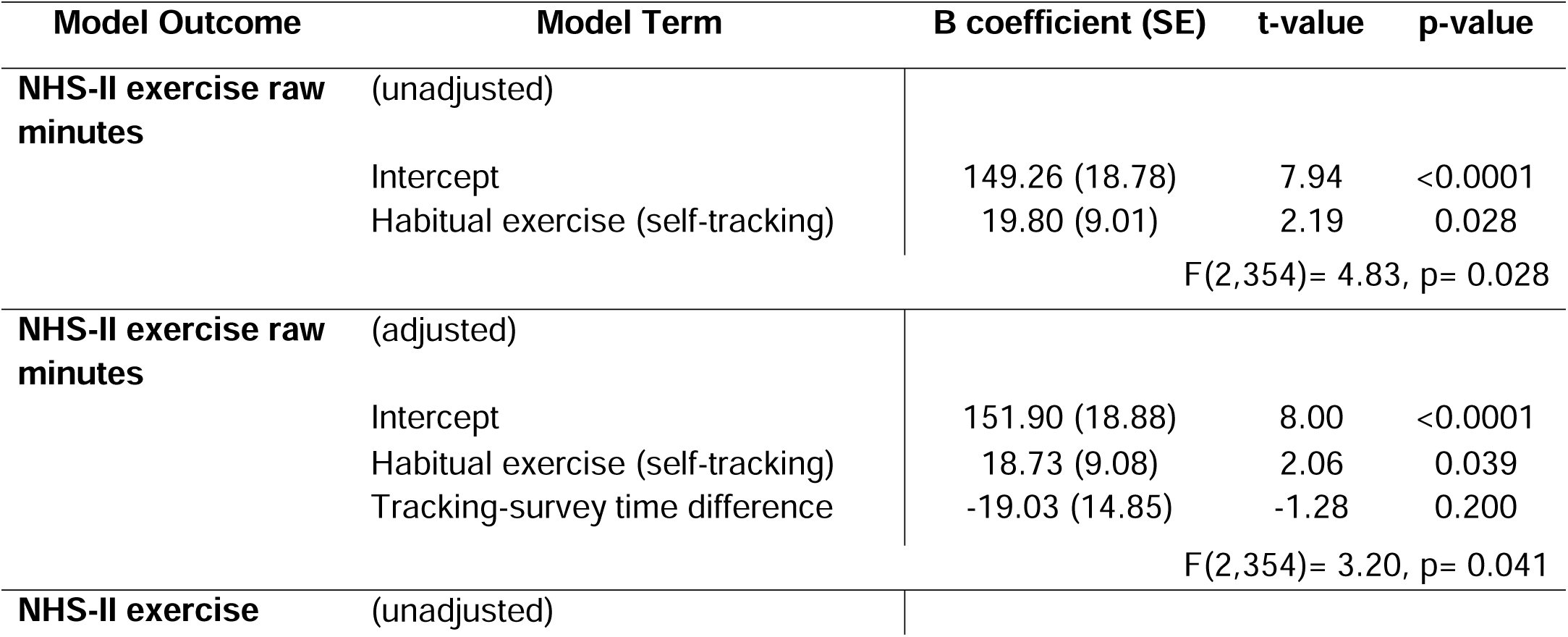
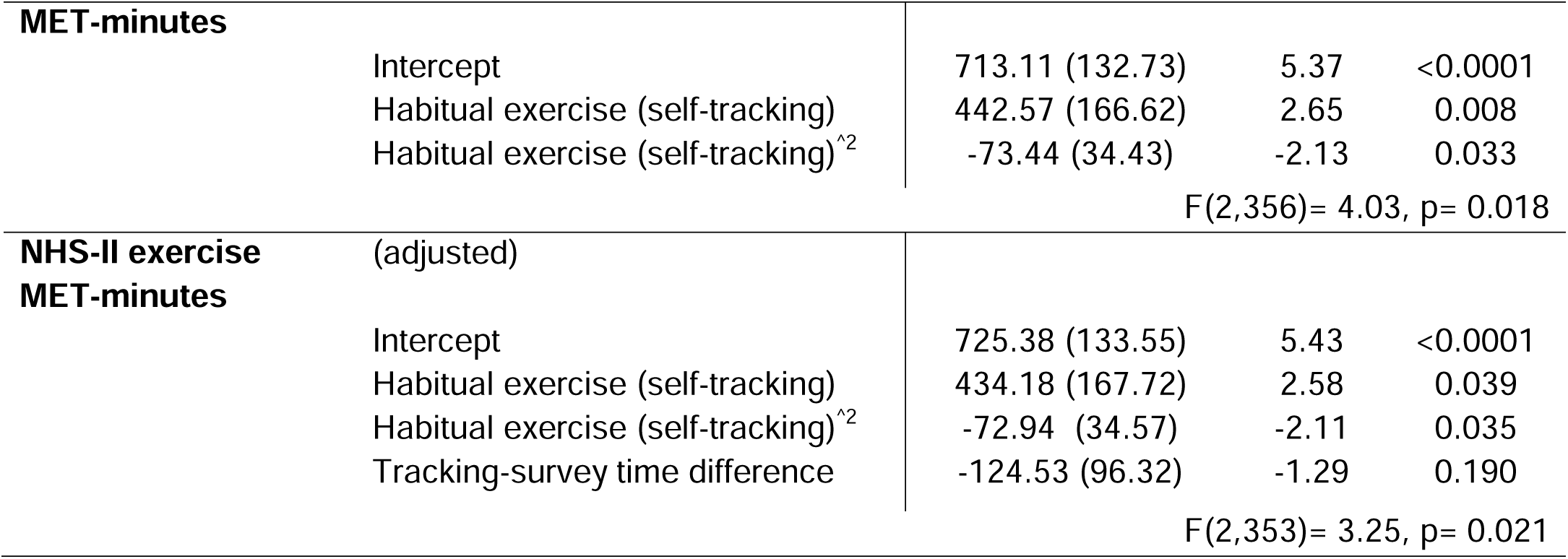
Results for the unadjusted and adjusted linear regression models estimating NHS-II weekly exercise minutes from median weekly exercise frequency scores from self-tracking Phendo.

There was a non-linear relationship between NHS-II weekly MET-mins of exercise and self-tracking based habitual exercise. The bivariate correlations indicated small-to-moderate associations (*τ* =0.27, z = 4.39, p<0.0001; corresponding to a Fisher’s z-score of 0.27). A 2° polynomial regression model provided the best fit for describing the association where, each self-tracked exercise instance in a typical week was associated with an additional ∼ 434-442 MET-mins on the NHS-II (B=442.57 and B=434.18 for the unadjusted and adjusted models, respectively). Moreover, these effects were independent of the time difference between the self-tracking period and survey completion, based on the associated non-significant point estimate (See Table 6).

### Discriminant Validity

Results of the known-group differences analysis of habitual exercise based on SF-36 subscales and BMI, along with mean scores on grouping variables and habitual exercise point estimates are provided in Table 7. Those with above population normative scores (i.e., >50) on the SF-36 Physical Function and Energy subscales had significantly lower levels of habitual exercise compared to those who had scores below the norm (t=-2.19, p=0.029, t=-2.75, p=0.008, respectively). For BMI category, those who had a BMI of 30 or higher were associated with significantly lower levels of habitual exercise compared to those with a BMI within the healthy range (i.e., 18.5-29.99).

**Table 7.** Results of the t-tests and linear regressions comparing group differences in habitual exercise levels.

| Grouping Variable (N) | Mean (SD) | Model point estimate (habitual exercise) |
| --- | --- | --- |
| <u>SF-36 Physical Function</u> |  | 1.56 |
| >50 (257) | 33.7 (14.7) | 1.20 |
| <=50 (117) | 78.9 (14.4) |  |
| | | $t=-2.19$ (228.59), $p=0.029$ |
| <u>SF-36 Energy</u> |  |  |
| >50 (339) | 64.6 (8.29) | 1.37 |
| <=50 (34) | 22.1 (13.9) | 2.16 |
| $t=-2.75 (38.89), p=0.008$ | | |
| <u>BMI</u> |  |  |
| <18.5 (19) | 17.6 (0.91) | 0.48 |
| 18.5-24.99 (146) | 21.9 (1.67) | 1.49 |
| 25-29.99 (59) | 26.7 (1.38) | 0.16 |
| >=30 (60) | 36.1 (5.09) | -0.45 |
| $F(3,280)=2.77, p=0.04$ | | |

## Discussion

### Overall summary of findings

This study provides the first line of evidence toward validation of a brief and customizable mHealth-based measure for self-tracking exercise designed for individuals with endometriosis. The results suggest that responses from Phendo’s exercise measure are moderately congruent with those from other self-reported recall-based and objectively-estimated exercise outcomes. Moreover, our preliminary results indicate that the self-tracking item can be used to assess different time frames (i.e., day-level and longitudinal). These findings collectively provide promising evidence that a simple and brief digital measure might be a sufficient tool for assessing exercise behavior in individuals with endometriosis, and potentially other women’s reproductive conditions.

### Day-level overall exercise concurrent validity

In Study 1, responses from the exercise self-tracking item when used as an overall (i.e., binary) measure of day-level exercise were moderately correlated with minutes of MVE and tracker-based steps (i.e., *τ*=0.256 and *τ*=0.295, respectively; corresponding to Fisher’s Z scores of 0.26 and 0.30). Moreover, each self-tracked exercise instance was associated with ∼18 minutes of MVE based on the point estimates in the adjusted and unadjusted regression models. These findings indicate that as a measure of overall day-level exercise, our brief self-tracking item has acceptable concurrent validity. In addition, we provide evidence that each self-tracked instance is predictive of an ∼18 minute bout of MVE, which can be useful when there is a need for a brief and simple measure that could be used to determine whether the participant meets the PA guidelines.

### Day-level aerobic and anaerobic/multimodal exercise validity

As expected, congruency was higher when the self-tracking item responses were limited to aerobic-based (including walking) exercise modalities (i.e., Kendall’s *τ*=0.397 and *τ*=0.361, respectively). On the other hand, self-tracked multimodal exercise responses were weakly correlated and not statistically significant. Results of the regression analyses further supported these findings, where the self-tracked aerobic exercise responses were associated with stronger point estimates. Specifically, each self-tracked aerobic exercise instance was associated with ∼28 mins of MVE and ∼3,924 steps on a given day. These findings are in line with our hypotheses and provide preliminary evidence toward the concurrent validity of the free-text responses from the custom-created self-tracking items. This has important implications from a researcher’s perspective, as the ability to use items that can be tailored based on participant preferences and needs can have benefits (e.g., engaging/retaining participants, ensuring the relevance of the outcomes collected). While these findings are promising; there is nevertheless a need to investigate best practices for evaluation of mHealth measures designed for event-based self-tracking and those that are customizable. To date, there have been few studies evaluating similar mHealth measures for self-monitoring[9, 10] and those that can be tailored,[48] but none that focus on PA and exercise behavior that are customizable by the user and allow daily assessments. Given the population prevalence of physical inactivity and its public health implications, this would be an important point of inquiry for future studies.

### Habitual exercise measure concurrent validity

The results from Study 2 indicated that the self-tracking based habitual exercise scores were predictive of NHS-II leisure-time PA (i.e., exercise) scores. For the average participant, each additional self-tracked exercise frequency in a typical week corresponded to 18.7 raw minutes on the NHS-II scale, independent of the time duration between self-tracking period and NHS-II survey completion. This is in line with our findings from the regression analyses in Study 1, where each self-tracked any exercise was associated with 16.1 minutes of MVE. Similarly, for the average participant, each additional self-tracked exercise frequency in a typical week corresponded to ∼434 MET-mins of exercise. To put this in perspective, a brisk walk at 3 or 4 miles per hour corresponds to 240 MET-mins (i.e., based on an intensity of 4 METs assigned in the Compendium of PA). Jumping rope, as an example of more vigorous activity, corresponds to 738 MET-mins (MET = 12.3). Nevertheless, we refrain from making conclusive remarks regarding a possible one-to-one mapping between these 2 outcomes. The habitual exercise measure was derived using all self-tracked exercise responses and therefore is not selective of intensity or energy expenditure of exercise. Future studies are warranted to investigate the seemingly non-linear relationship between these 2 outcomes and to assess a habitual exercise outcome derived using only certain exercise modalities based on intensity, similar to Study 1.

### Habitual exercise measure discriminant validity

Results of the discriminant validity analysis in Study 2 indicated that habitual exercise levels significantly differed between individuals based on their SF-36 Physical Function and Energy sores, and BMI category. As expected, those who reported higher scores (i.e., indicating higher functioning and energy) and a BMI of <30 were associated with greater habitual exercise levels. It makes sense that individuals who experience greater physical dysfunction and fatigue would be less likely to engage in exercise or any other type of daily PA over the long term. Of note, those in the below-norm categories had group mean scores that were 2 and 3 SDs below the population means (i.e., 33.7 for Physical Function and 22.1 for Energy) in our sample. These findings are in line with previous studies that assess endometriosis-related impairment in daily functioning using mHealth-based self-tracking data.[5] Moreover, those in the 30+ BMI category (currently considered as “obese”) were statistically significantly less likely to report any exercise, based on the sample median habitual exercise frequency of zero. However, there were no significant differences between the other BMI categories. This finding is in line with our hypothesis and previous studies reporting significant differences in PA levels among those with a BMI of 30+ vs <30 using minute-by-minute accelerometry-estimated PA data.[47] In sum, our analyses provide preliminary evidence toward the discriminant ability of this habitual exercise measure and we note the convenience sample based selection of factors for conducting the known-group differences. To further ascertain its discriminant properties, further investigations are needed where a wider set of factors that are known to differ across varying levels of habitual exercise are used.

### Overall Implications

Overall, the results from this study suggest that a simple and brief exercise measure might be a sufficient tool for assessing exercise behavior at the day-level and for estimating one’s habitual exercise (i.e., patterns observed over the long term). This has useful implications for conducting large-scale studies that involve monitoring a diverse group of participants over long durations of time, as well as maintaining engagement and retention. In these scenarios, the ability to administer a brief measure that can further be customized based on individual needs and preferences can be advantageous. Similarly, it can be useful when faced with a variety of perceived or external barriers to self-monitoring (e.g., lack of time, technological limitations, varying literacy or interest in mHealth technology) that are applicable in both research and non-research setting. Specifically in the context of endometriosis, the ability to self-monitor, reflect on personal history of exercise behavior, and the customizability of the measure could all serve as contributing factors to increasing one’s exercise levels and self-efficacy. This is based on previous research indicating interest among individuals with endometriosis in self-monitoring symptoms and self-management behaviors over time toward finding solutions to better managing their condition. Nevertheless, future studies are warranted to investigate the relationship of self-tracking behavior to self-efficacy and whether it leads to increases in PA and/or exercise behavior. There are currently no validated self-report measures that are designed for frequent, repeated assessments of PA or exercise in endometriosis. Existing measures are based on recall of an extended past time period (e.g., previous week, month, week), and include multiple items. As such, they are not designed for daily use and such an attempt can be burdensome on the participant. Moreover, they do not allow customization based on participant’s needs and preferences. To this end, our findings constitute novel and important findings to the body of literature on PA and exercise measurement using self-report.

### Limitations

We acknowledge several limitations of this study. First, the study sample consisted primarily of White, non-Hispanic women with sufficient English comprehension for providing informed consent and using the App. As such, these results might not be generalizable to other demographic groups. Similarly, Study 1 included a relatively small sample size and the data from the activity trackers were based on self-report, as well as use of a variety of trackers by the participants. Thus, future studies are needed to investigate the association of the scores from Phendo’s self-tracking item to objectively-estimated measures in larger samples using the same accelerometers for all participants, and possibly for longer than 2 weeks. Another limitation was the variable time points at which the participants completed the NHS-II in Study 2. This resulted in a range of time differences between self-tracking in Phendo and when the participants completed the NHS-II Scale. We adjusted the regression models to account for this potentially confounding variable, however; this design can be improved by administering the surveys to all the participants at the same time point in relation to their self-tracking period (e.g., at the beginning or at the end). Similarly, we compared the habitual exercise measure to only 1 other scale, and we did not have other PA or exercise measures for comparison. Future studies can more comprehensively evaluate by implementing a nomological network approach where its associations with several measures of both convergent or divergent constructs are assessed.[49, 50]

## Conclusion

This study presents preliminary evidence on the concurrent and discriminant validity of a brief mHealth App measure for exercise self-tracking among individuals with endometriosis. Results suggest that a simple and brief digital measure might be a sufficient tool for assessing exercise in individuals with endometriosis, and potentially those with other women’s reproductive conditions. These findings have important implications in the context of large-scale studies that involve monitoring a diverse group of participants over long durations of time, reaching populations that might be harder to reach, as well as engaging and retaining research participants.

## Data Availability

The data produced in the present study are available upon reasonable request to the authors.

## Author Contributions

IE conceptualized the study, conducted the data analyses, and prepared the first draft of the manuscript. SB provided guidance on the study design and data analyses. ENH was responsible for data acquisition, curation and management. NE acquired the funding and provided the mHealth infrastructure for the study (Phendo App). SB, NE, and ENH reviewed and provided feedback on the manuscript.

## Data availability statement

Data are available on reasonable request.

## References

1. van Velthoven MH, Car J, Zhang Y, Marušić A. mHealth series: New ideas for mHealth data collection implementation in low- and middle-income countries. J Glob Health. 2013;3(2):020101-. doi: 10.7189/jogh.03.020101. PubMed PMID: 24363911.

2. Pham Q, Wiljer D, Cafazzo JA. Beyond the Randomized Controlled Trial: A Review of Alternatives in mHealth Clinical Trial Methods. JMIR Mhealth Uhealth. 2016;4(3):e107. doi: 10.2196/mhealth.5720.

3. Epstein DH, Tyburski M, Kowalczyk WJ, Burgess-Hull AJ, Phillips KA, Curtis BL, et al. Prediction of stress and drug craving ninety minutes in the future with passively collected GPS data. NPJ digital medicine. 2020;3:26-. doi: 10.1038/s41746-020-0234-6. PubMed PMID: 32195362.

4. Adams P, Murnane EL, Elfenbein M, Wethington E, Gay G, editors. Supporting the self-management of chronic pain conditions with tailored momentary self-assessments. Proceedings of the 2017 CHI Conference on Human Factors in Computing Systems; 2017.

5. Ensari I, Pichon A, Lipsky-Gorman S, Bakken S, Elhadad N. Augmenting the Clinical Data Sources for Enigmatic Diseases: A Cross-Sectional Study of Self-Tracking Data and Clinical Documentation in Endometriosis. Applied Clinical Informatics. 2020;11(05):769–84.

6. Schneider S, Stone AA, Schwartz JE, Broderick JE. Peak and end effects in patients’ daily recall of pain and fatigue: a within-subjects analysis. The journal of pain. 2011;12(2):228–35. doi: 10.1016/j.jpain.2010.07.001. PubMed PMID: 20817615.

7. Shiffman S, Stone AA, Hufford MR. Ecological momentary assessment. Annu Rev Clin Psychol. 2008;4:1-32. Epub 2008/05/30. doi: 10.1146/annurev.clinpsy.3.022806.091415. PubMed PMID: 18509902.

8. Smyth JM, Stone AA. Ecological Momentary Assessment Research in Behavioral medicine. Journal of Happiness Studies. 2003;4(1):35–52. doi: 10.1023/A:1023657221954.

9. Swendeman D, Comulada WS, Ramanathan N, Lazar M, Estrin D. Reliability and Validity of Daily Self-Monitoring by Smartphone Application for Health-Related Quality-of-Life, Antiretroviral Adherence, Substance Use, and Sexual Behaviors Among People Living with HIV. AIDS and Behavior. 2015;19(2):330–40. doi: 10.1007/s10461-014-0923-8.

10. Swendeman D, Comulada WS, Koussa M, Worthman CM, Estrin D, Rotheram-Borus MJ, et al. Longitudinal Validity and Reliability of Brief Smartphone Self-Monitoring of Diet, Stress, and Physical Activity in a Diverse Sample of Mothers. JMIR Mhealth Uhealth. 2018;6(9):e176. doi: 10.2196/mhealth.9378.

11. Aung MSH, Alquaddoomi F, Hsieh C-K, Rabbi M, Yang L, Pollak JP, et al. Leveraging multi-modal sensing for mobile health: a case review in chronic pain. IEEE journal of selected topics in signal processing. 2016;10(5):962–74.

12. Kumar S, Nilsen WJ, Abernethy A, Atienza A, Patrick K, Pavel M, et al. Mobile health technology evaluation: the mHealth evidence workshop. Am J Prev Med. 2013;45(2):228–36. doi: 10.1016/j.amepre.2013.03.017. PubMed PMID: 23867031.

13. McKillop M, Voigt N, Schnall R, Elhadad N. Exploring self-tracking as a participatory research activity among women with endometriosis. Journal of Participatory Medicine. 2016.

14. McKillop M, Mamykina L, Elhadad N, editors. Designing in the Dark: Eliciting Self-tracking Dimensions for Understanding Enigmatic Disease. Proceedings of the 2018 CHI Conference on Human Factors in Computing Systems; 2018: ACM.

15. Fourquet J, Gao X, Zavala D, Orengo JC, Abac S, Ruiz A, et al. Patients’ report on how endometriosis affects health, work, and daily life. Fertil Steril. 2010;93(7):2424–8. doi: 10.1016/j.fertnstert.2009.09.017. PubMed PMID: 19926084.

16. Schliep KC, Mumford SL, Peterson CM, Chen Z, Johnstone EB, Sharp HT, et al. Pain typology and incident endometriosis. Hum Reprod. 2015;30(10):2427-38. Epub 2015/08/11. doi: 10.1093/humrep/dev147. PubMed PMID: 26269529.

17. De Graaff A, D’hooghe T, Dunselman G, Dirksen C, Hummelshoj L, Consortium WE, et al. The significant effect of endometriosis on physical, mental and social wellbeing: results from an international cross-sectional survey. Human reproduction. 2013;28(10):2677–85.

18. Simoens S, Dunselman G, Dirksen C, Hummelshoj L, Bokor A, Brandes I, et al. The burden of endometriosis: costs and quality of life of women with endometriosis and treated in referral centres. Human Reproduction. 2012;27(5):1292–9.

19. Soliman AM, Coyne KS, Gries KS, Castelli-Haley J, Snabes MC, Surrey ES. The effect of endometriosis symptoms on absenteeism and presenteeism in the workplace and at home. Journal of managed care & specialty pharmacy. 2017;23(7):745–54.

20. Simoens S, Dunselman G, Dirksen C, Hummelshoj L, Bokor A, Brandes I, et al. The burden of endometriosis: costs and quality of life of women with endometriosis and treated in referral centres. Hum Reprod. 2012;27(5):1292-9. Epub 2012/03/17. doi: 10.1093/humrep/des073. PubMed PMID: 22422778.

21. Pokrzywinski RM, Soliman AM, Chen J, Snabes MC, Coyne KS, Surrey ES, et al. Achieving clinically meaningful response in endometriosis pain symptoms is associated with improvements in health-related quality of life and work productivity: analysis of 2 phase III clinical trials. American Journal of Obstetrics and Gynecology. 2020;222(6):592.e1-e10. doi: https://doi.org/10.1016/j.ajog.2019.11.1255.

22. The Practice Committee of the American Society for Reproductive Medicine. Treatment of pelvic pain associated with endometriosis: a committee opinion. 2014 2014/04/01/. Report No.: 0015-0282 Contract No.: 4.

23. Ambrose KR, Golightly YM. Physical exercise as non-pharmacological treatment of chronic pain: Why and when. Best Pract Res Clin Rheumatol. 2015;29(1):120-30. Epub 2015/05/23. doi: 10.1016/j.berh.2015.04.022. PubMed PMID: 26267006.

24. Rice D, Nijs J, Kosek E, Wideman T, Hasenbring MI, Koltyn K, et al. Exercise-Induced Hypoalgesia in Pain-Free and Chronic Pain Populations: State of the Art and Future Directions. The Journal of Pain. 2019;20(11):1249–66. doi: https://doi.org/10.1016/j.jpain.2019.03.005.

25. Lemmens J, De Pauw J, Van Soom T, Michiels S, Versijpt J, van Breda E, et al. The effect of aerobic exercise on the number of migraine days, duration and pain intensity in migraine: a systematic literature review and meta-analysis. J Headache Pain. 2019;20(1):16. Epub 2019/02/16. doi: 10.1186/s10194-019-0961-8. PubMed PMID: 30764753; PubMed Central PMCID: PMCPMC6734345.

26. Armour M, Ee CC, Naidoo D, Ayati Z, Chalmers KJ, Steel KA, et al. Exercise for dysmenorrhoea. Cochrane Database of Systematic Reviews. 2019;(9). doi: 10.1002/14651858.CD004142.pub4. PubMed PMID: CD004142.

27. Rabbi M, Aung MS, Gay G, Reid MC, Choudhury T. Feasibility and acceptability of mobile phone–based auto-personalized physical activity recommendations for chronic pain self-management: pilot study on adults. Journal of medical Internet research. 2018;20(10):e10147.

28. Sevel L, Boissoneault J, Alappattu M, Bishop M, Robinson M. Training endogenous pain modulation: a preliminary investigation of neural adaptation following repeated exposure to clinically-relevant pain. Brain Imaging and Behavior. 2020;14(3):881–96. doi: 10.1007/s11682-018-0033-8.

29. Gordon R, Bloxham S, editors. A systematic review of the effects of exercise and physical activity on non-specific chronic low back pain. Healthcare; 2016: Multidisciplinary Digital Publishing Institute.

30. Stain HJ, Bjornestad J. Chapter 9 - Assessing Social Functioning Across the Life Course of Psychosis. In: Badcock JC, Paulik G, editors. A Clinical Introduction to Psychosis: Academic Press; 2020. p. 207–22.

31. Krabbe PFM. Chapter 7 - Validity. In: Krabbe PFM, editor. The Measurement of Health and Health Status. San Diego: Academic Press; 2017. p. 113–34.

32. Frost MH, Reeve BB, Liepa AM, Stauffer JW, Hays RD. What is sufficient evidence for the reliability and validity of patient-reported outcome measures? Value Health. 2007;10 Suppl 2:S94-s105. Epub 2007/11/13. doi: 10.1111/j.1524-4733.2007.00272.x. PubMed PMID: 17995479.

33. Anthoine E, Moret L, Regnault A, Sébille V, Hardouin J-B. Sample size used to validate a scale: a review of publications on newly-developed patient reported outcomes measures. Health Qual Life Outcomes. 2014;12:176-. doi: 10.1186/s12955-014-0176-2. PubMed PMID: 25492701.

34. US Department of Health Human Services. Guidance for industry-Patient-reported outcome measures: Use in medical product development to support labeling claims. 2009.

35. Lomas J, Pickard L, Mohide A. Patient versus clinician item generation for quality-of-life measures: the case of language-disabled adults. Medical Care. 1987:764–9.

36. Ainsworth BE, Haskell WL, Herrmann SD, Meckes N, Bassett DR, Tudor-Locke C, et al. 2011 Compendium of Physical Activities: a second update of codes and MET values. Med Sci Sports Exerc. 2011;43(8):1575–81.

37. Coleman KJ, Ngor E, Reynolds K, Quinn VP, Koebnick C, Young DR, et al. Initial validation of an exercise “vital sign” in electronic medical records. Med Sci Sports Exerc. 2012;44(11):2071-6. Epub 2012/06/13. doi: 10.1249/MSS.0b013e3182630ec1. PubMed PMID: 22688832.

38. Ensari I, Gorman S, Horan E, Bakken S, Elhadad N. Characterizing Associations of Exercise and Pain Patterns in Endometriosis via Mobile Self-Tracking The Journal of Pain. 2021.

39. Wolf AM, Hunter DJ, Colditz GA, Manson JE, Stampfer MJ, Corsano KA, et al. Reproducibility and validity of a self-administered physical activity questionnaire. Int J Epidemiol. 1994;23(5):991-9. Epub 1994/10/01. doi: 10.1093/ije/23.5.991. PubMed PMID: 7860180.

40. Vitonis AF, Vincent K, Rahmioglu N, Fassbender A, Buck Louis GM, Hummelshoj L, et al. World Endometriosis Research Foundation Endometriosis Phenome and Biobanking Harmonization Project: II. Clinical and covariate phenotype data collection in endometriosis research. Fertility and sterility. 2014;102(5):1223-32. Epub 2014/09/22. doi: 10.1016/j.fertnstert.2014.07.1244. PubMed PMID: 25256930.

41. Jones G, Kennedy S, Barnard A, Wong J, Jenkinson C. Development of an endometriosis quality-of-life instrument: The Endometriosis Health Profile-30. Obstet Gynecol. 2001;98(2):258-64. Epub 2001/08/17. doi: 10.1016/s0029-7844(01)01433-8. PubMed PMID: 11506842.

42. Dencker M, Andersen LB. Accelerometer-measured daily physical activity related to aerobic fitness in children and adolescents. Journal of Sports Sciences. 2011;29(9):887–95. doi: 10.1080/02640414.2011.578148.

43. Hills AP, Mokhtar N, Byrne NM. Assessment of Physical Activity and Energy Expenditure: An Overview of Objective Measures. Frontiers in Nutrition. 2014;1. doi: 10.3389/fnut.2014.00005.

44. McHorney CA, Ware JE, Lu JFR, Sherbourne CD. The MOS 36-Item Short-Form Health Survey (SF-36): III. Tests of Data Quality, Scaling Assumptions, and Reliability across Diverse Patient Groups. Medical Care. 1994;32(1):40–66.

45. Hays RD, Sherbourne CD, Mazel R. User’s Manual for the Medical Outcomes Study (MOS) Core Measures of Health-Related Quality of Life. Santa Monica, CA: RAND Corporation; 1995.

46. Bakken S, Grullon-Figueroa L, Izquierdo R, Lee N-J, Morin P, Palmas W, et al. Development, validation, and use of English and Spanish versions of the telemedicine satisfaction and usefulness questionnaire. J Am Med Inform Assoc. 2006;13(6):660-7. Epub 2006/08/23. doi: 10.1197/jamia.M2146. PubMed PMID: 16929036.

47. Cooper AR, Page A, Fox KR, Misson J. Physical activity patterns in normal, overweight and obese individuals using minute-by-minute accelerometry. Eur J Clin Nutr. 2000;54(12):887-94. Epub 2000/01/11. doi: 10.1038/sj.ejcn.1601116. PubMed PMID: 11114687.

48. Ruland CM, Bakken S, Røislien J. Reliability and Validity Issues Related to Interactive Tailored Patient Assessments: A Case Study. J Med Internet Res. 2007;9(3):e22. doi: 10.2196/jmir.9.3.e22.

49. Ensari I, Motl RW, McAuley E. Structural and construct validity of the Leeds Multiple Sclerosis Quality of Life scale. Quality of Life Research. 2016;25(6):1605–11.

50. Cronbach LJ, Meehl PE. Construct validity in psychological tests. Psychological bulletin. 1955;52(4):281.

